# Cannabidiol blood metabolite levels after cannabidiol treatment are associated with broadband EEG changes and improvements in visuomotor and non-verbal cognitive abilities in boys with autism requiring higher levels of support

**DOI:** 10.1101/2024.09.27.24314448

**Authors:** Christian Cazares, Austin Hutton, Gisselle Paez, Doris Trauner, Bradley Voytek

## Abstract

Oral cannabidiol (CBD) treatment has been suggested to alleviate severe symptoms of autism spectrum disorder (ASD). While many CBD preparations have been studied in clinical trials involving ASD, none has used purified CBD preparations or preparations approved by the U.S. Food and Drug Administration, nor have they focused on children with ASD with higher support needs. Previous studies have identified several candidate electrophysiological biomarkers of cognitive and behavioral disabilities in ASD, with emerging biomarkers including periodic (oscillatory) and aperiodic measures of neural activity. We analyzed electroencephalography (EEG) recordings from 24 boys with ASD and higher support needs (aged 7-14 years) from a prior double-blind, placebo-controlled, crossover Phase II Clinical Trial (NCT04517799) that investigated whether 8 weeks of daily CBD treatment (titrated to 20 mg/kg/day) improved severe behavioral problems, measured at baseline, post-CBD, post-placebo, and post-washout. Using linear mixed effect models, we found that aperiodic EEG measures varied with CBD metabolite levels in blood, as evidenced by a larger aperiodic offset across the scalp and a decreased aperiodic exponent across occipital electrodes. Furthermore, CBD metabolite levels in blood had a positive association with receptive vocabulary, nonverbal intelligence and visuomotor coordination. Our data suggest that this daily CBD preparation and administration schedule produced mixed effects, with some children showing improvements in cognitive and behavioral abilities while others demonstrated limited changes. Our findings support the inclusion of aperiodic EEG measures alongside traditional oscillatory EEG measures as candidate biomarkers for tracking the variable clinical impact of purified CBD treatment in children with ASD.

## 1. Introduction

Neurodevelopmental disorders (NDDs), including autism spectrum disorder (ASD), may be characterized by severe cognitive and behavioral dysfunction that impose significant burdens on caregivers, families, and finances [1]. The core symptoms of ASD include deficits in social communication and stereotyped repetitive behaviors, but other co-occurring behaviors such as self-injurious and aggressive behaviors [2], may require intervention. Current pharmacotherapies have limited efficacy, and progress in developing new treatments is hindered by a lack of reliable biomarkers that can identify patients who are most likely to benefit from investigational therapies in early life [3].

Local field potential signals, such as those recorded by scalp electroencephalography (EEG), comprise mixed periodic and aperiodic components. These components are thought to reflect synchronized and asynchronous neuronal firing in cortical networks, respectively, and can serve as putative indices of cortical excitation-inhibition (E:I) balance in health and disease [4–9]. These components correlate with cognitive processes and show alterations in a wide variety of conditions associated with E:I imbalance, including NDDs [7,10–15]. Task-free EEG has proven valuable for investigating neural correlates in intellectually impaired and minimally verbal pediatric populations [10,14,16,17]. EEG abnormalities in NDDs likely result from impaired neuronal maturation during early development [18–20]. Previous research has largely focused on periodic components, such as the oscillatory sensorimotor mu rhythm and visual cortical alpha rhythms, both in the ∼8-12 Hz frequency range [21–23]. Peak alpha frequency (6-12 Hz) has been identified as a biomarker for non-verbal cognition in ASD [24], while delta rhythms (1-4 Hz) show disruptions in ASD [16,25]. Building on these findings, identifying complementary aperiodic EEG signal features that are thought to reflect cortical E:I balance could further inform the development of pharmacotherapies and biomarkers for severe ASD.

Medications like risperidone and aripiprazole show efficacy in improving ASD behavioral symptoms [26], but their adverse effects require close medical monitoring [27]. This has led to growing interest in cannabidiol (CBD), a non-psychoactive cannabis-derived compound that has shown promising anecdotal and clinical trial outcomes for children with intractable epilepsy and co-morbid ASD, with mild side effects [28–32]. This led to U.S. Food and Drug Administration (FDA) approval of Epidiolex®, a plant-based purified CBD medication for treatment of intractable childhood epilepsies. During the same time frame, there have been a number of investigations relating to the use of non-FDA-approved CBD-based medications in other conditions, including retrospective studies in children with ASD, where participants exhibited improvements in disruptive behavior when treated with CBD:THC (20:1) formulations [33], with parents reporting reduced aggression, hyperactivity and anxiety [34]. Social communication improvements have also been reported following 6 months of CBD:THC formulation in children and adolescents with ASD [35,36], with speculation that these improvements are mediated by oxytocin-dependent endocannabinoid signaling as shown in rodents [37,38]. If behavioral and social improvements are mediated through the endocannabinoid system, it raises the question whether purified CBD can lead to similar improvements without the need for a psychoactive THC component in its formulation. Recent evidence of clinical improvement with CBD comes from a clinical trial in which blinded clinicians observed reductions in aggressive behaviors and hyperactivity, as well as improvements in communication [39].

Single doses of CBD modulate GABA levels in prefrontal regions [40], potentially affecting cortical E:I balance through GABAergic inhibition, which has been proposed to be assessed from spectral parameterization of EEG signals [41–43]. Although CBD hasn’t been directly studied in ASD animal models, studies in a Dravet syndrome mouse model showed improvements in social deficits [44], and CBD administration in adolescent mice yielded modest spatial memory improvements without negative impacts [45]. CBD-based medication is thus a viable candidate for investigating therapeutic effects on cognitive and behavioral symptoms in children with ASD, given its regulatory precedent, promising safety profile, and preliminary data from both human and mouse models. However, there has been no longitudinal, placebo-controlled monitoring of brain activity and cognitive-behavioral abilities in children with ASD requiring higher levels of support receiving CBD treatment.

Here we investigated periodic and aperiodic measures of EEG signals as potential biomarkers for CBD treatment effects in ASD. We hypothesized that EEG signal measures and cognitive-behavioral outcomes would be associated with CBD treatment responses, as indicated by active and inactive CBD metabolite levels in blood before and after an 8-week oral CBD regimen in boys with ASD and higher support needs [46–48].

## 2. Patients and Methods

Below is a condensed version of Patients and Methods. A detailed version can be found in the Supplementary Materials.

### 2.1. Experimental recordings and cognitive-behavioral testing

#### 2.1.1. Participants

Our data was sourced from a clinical trial performed at the University of California, San Diego [39]. The trial was approved by the institutional review board of the University of California, San Diego and registered with http://www.clinicaltrials.gov (NCT04517799) to investigate Epidiolex® (CBD) effects on severe behavioral issues in boys with ASD. The dataset included in this study consisted of 24 males (ages 7-14) who had prior clinical diagnoses of ASD using DSM-5 criteria.

The diagnoses were made by clinicians with expertise and experience in diagnosing and treating children with ASD. All participants underwent Autism Diagnostic Observation Schedule, Second Edition (ADOS-2) assessments of symptoms and exhibited severe behavioral issues including stereotypies, aggression, self-injury, and/or hyperactivity. Baseline cognitive and behavioral assessments (Table 1) revealed the extent of impairments, with many participants unable to attempt certain standardized assessments or performing below normative ranges, supporting their characterization as having higher support needs. All participants were reported to require continuous supervision of activities of daily living and personal safety. The participants were free of other neurological conditions, including epilepsy. Parental consent was obtained, with child assent waived due to participants’ cognitive limitations.

**Table 1.** Baseline participant characteristics. Raw score and standard score means with standard error of the mean (SEM), ranges, and floor effects for cognitive and behavioral assessments at baseline. Raw Score N indicates participants who attempted each assessment, while Standard Score N indicates participants who achieved valid standard scores, with the gap between these values reflecting performance below normative ranges.

|  | Raw Score N | Raw Score Mean (SEM) | Raw Score Range | Standard Score N | Standard Score Mean (SEM) | Standard Score Range | Floor Effects n(%) |
| --- | --- | --- | --- | --- | --- | --- | --- |
| PPVT-4 (Receptive Vocabulary) | 17 | 95.8 (14.7) | 17 to 204 | 17 | 64.1 (6.2) | 40 to 115 | 5 (29%) |
| TONI-4 (Nonverbal Intelligence) | 14 | 19.3 (3.4) | 2 to 37 | 14 | 86.8 (4.6) | 61 to 121 | 0 (0%) |
| EOWPVT-4 (Expressive Vocabulary) | 13 | 59.5 (11.8) | 18 to 146 | 6 | 87.5 (9.0) | 63 to 124 | 0 (0%) |
| Beery VMI (Visual-Motor Integration) | 21 | 12.9 (1.6) | 0 to 30 | 14 | 67.1 (5.8) | 45 to 112 | 1 (7%) |
| Beery VP (Visual Perception) | 21 | 9.6 (2.2) | 0 to 29 | 7 | 88.1 (7.2) | 62 to 118 | 0 (0%) |
| Beery MC (Motor Coordination) | 21 | 11.4 (1.6) | 1 to 30 | 10 | 64.1 (6.3) | 45 to 109 | 2 (20%) |
| RBS-R Total (Repetitive Behaviors) | 21 | 59.7 (4.6) | 24 to 93 | - | - | - | - |
| ADOS-2 Module | 21 (1: 12, 2: 3, 3: 6) | - | - | - | - | - | - |

#### 2.1.2 Experimental design

Our dataset consisted of assessments made via a randomized, two-arm design (CBD-Placebo or Placebo-CBD) with treatment periods lasting 8 weeks, separated by a 4-week washout period. Cognitive-behavioral assessments and task-free scalp EEG recordings were collected at baseline and after each timepoint (Figure 1A). CBD dosage was determined based on participant weight starting with 5 mg/kg/day, with a gradual increase over the first two weeks to reach the maximum dose (20 mg/kg/day, divided into two oral doses) for the remainder of the study period. Whole blood samples analyzed in this study were collected near EEG and cognitive-behavioral assessments. CBD metabolite levels were quantified alongside the major active metabolite 7-hydroxy-cannabidiol (7-OH-CBD) and inactive metabolite 7-carboxy-cannabidiol (7-COOH-CBD) [46,48,49]. Analysis of CBD metabolite concentrations in blood samples following the 8-week CBD treatment regimen revealed levels comparable to those found in a prior report using orally administered CBD (Epidiolex®) for four days in healthy adults [46] (Figure Supplement 1A-C).

**Figure 1.**
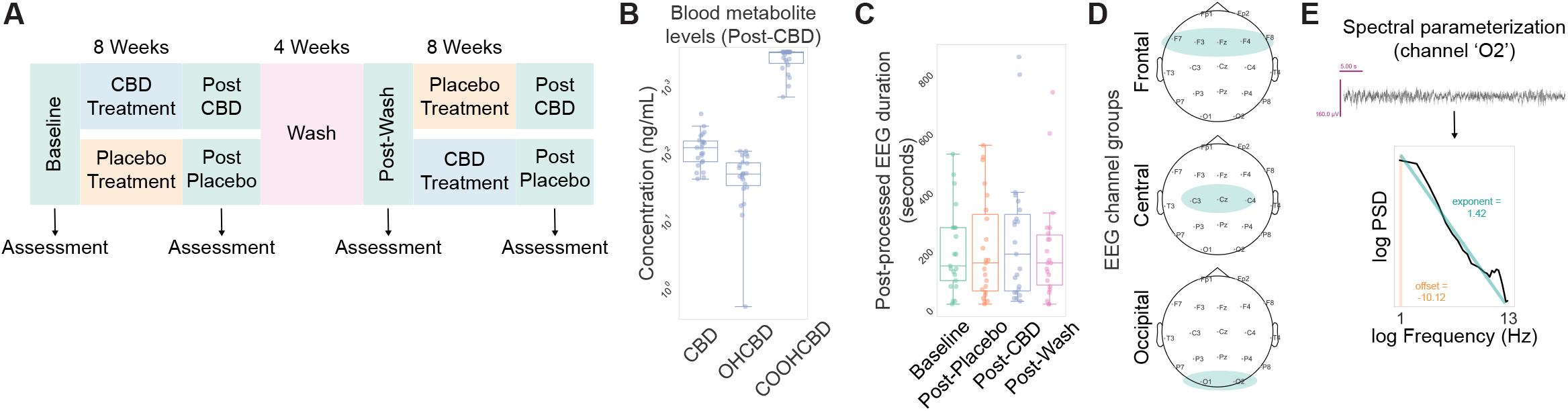
Overview of study design and spectral parameterization approach. **(A)** Schematic representation of data included in our analyses, comprising four study time points for EEG, blood draws, and cognitive-behavioral assessments. **(B)** Concentration of metabolite levels in blood post-CBD treatment. **(C)** Duration of post-processed EEG traces across study time points. **(D)** Schematic of electrodes and electrode groups used in this study. **(G)** Representative power spectrum of EEG channel ‘O2’ alongside extracted aperiodic exponent and offset values.

#### 2.1.3 Cognitive-behavioral testing

Assessments included Autism Diagnostic Observation Schedule, Second Edition (ADOS-2) Modules 1-3, Test of Nonverbal Intelligence, Fourth Edition (TONI-4) for nonverbal intelligence [50], Repetitive Behavior Scale-Revised (RBS-R) for repetitive behaviors [51], Peabody Picture Vocabulary Test, Fourth Edition (PPVT-4) for receptive vocabulary [52], Expressive One-Word Picture Vocabulary Test, Fourth Edition (EOWPVT-4) for expressive vocabulary [53], and Beery-Buktenica Developmental Test of Visual-Motor Integration, Sixth Edition (Beery VMI-6) for visual-motor integration, including supplemental subtests for visual perception (Beery VP) and motor coordination (Beery MC) [54]. Standard scores were calculated where possible for sample characterization purposes at baseline, while raw scores were used for model analyses to avoid floor effects and maintain sensitivity to change in this severely impaired population.

#### 2.1.4 EEG recording setup

EEG data were recorded using a CGX Quick-20 Dry wireless headset with 20 channels in 10-20 montage, online referenced to A1. Signals were sampled at 500 Hz (0-131 Hz bandwidth, 24-bit resolution), with electrode impedance maintained below 500 kΩ prior to the start of each recording session based on the manufacturer’s specifications. Task-free EEG recordings were made during awake, passive viewing periods with the goal of capturing 5 to 10 minutes during which the child remained calm by viewing a video of their choice.

### 2.2. Data analysis

#### 2.2.1. Preprocessing

Analysis was performed in Python using numpy [55], scipy [56], autoreject [57], MNE [58], specparam (formerly known as FOOOF) [59], and statsmodel [60]. EEG data was re-referenced offline to a linked ears reference (A1/A2) and preprocessed using Autoreject [57] to establish global artifact rejection thresholds. Data was bandpass filtered (0.1-100 Hz) and processed with ICA (“extended infomax”) to remove non-brain components per ICLabel criteria [61] (Figure Supplement 1B). Final analysis included 89 sessions across timepoints (Baseline: 21, Post-CBD: 23, Post-Placebo: 23, Post-Wash: 22), with 3-4 recordings per participant (Table 2).

**Table 2.** Participant assessments included in linear mixed effects models. Mean and standard error of the mean (SEM) for participant ages and assessment scores, alongside total number of participants (N) and total sessions included in each linear mixed model (n), across study time points.

|  | Baseline | Post-Placebo | Post-Wash | Post-CBD |
| --- | --- | --- | --- | --- |
| **Total participants (N)** | **21** | **23** | **22** | **23** |
| Age mean | 10.43 | 10.43 | 10.64 | 10.63 |
| Age SEM | 0.44 | 0.45 | 0.48 | 0.45 |
| EEG participants (n) | 21 | 23 | 22 | 23 |
| RBS-R participants (n) | 21 | 23 | 22 | 23 |
| RBS-R mean | 59.74 | 45.53 | 48.45 | 43.30 |
| RBS-R SEM | 4.57 | 3.67 | 3.70 | 3.81 |
| PPVT-4 participants (n) | 14 | 13 | 15 | 13 |
| PPVT-4 mean | 99.79 | 100.69 | 97.47 | 99.62 |
| PPVT-4 SEM | 15.50 | 16.87 | 15.81 | 17.81 |
| TONI-4 participants (n) | 12 | 13 | 14 | 13 |
| TONI-4 mean | 88.83 | 88.38 | 88.50 | 89.85 |
| TONI-4 SEM | 5.11 | 5.36 | 5.34 | 5.39 |
| EOWPVT-4 participants (n) | 13 | 13 | 12 | 12 |
| EOWPVT-4 mean | 59.54 | 57.92 | 57.17 | 61.50 |
| EOWPVT-4 SEM | 11.80 | 11.95 | 11.73 | 12.80 |
| Beery-VMI participants (n) | 21 | 23 | 22 | 21 |
| Beery-VMI mean | 12.90 | 13.87 | 14.45 | 13.57 |
| Beery-VMI SEM | 1.59 | 1.33 | 1.36 | 1.64 |
| Beery-VP participants (n) | 21 | 23 | 22 | 21 |
| Beery-VP mean | 9.62 | 11.04 | 12.32 | 9.90 |
| Beery-VP SEM | 2.19 | 2.27 | 2.27 | 2.29 |
| Beery-MC participants (n) | 21 | 23 | 22 | 21 |
| Beery-MC mean | 11.38 | 11.52 | 10.73 | 10.67 |
| Beery-MC SEM | 1.56 | 1.34 | 1.25 | 1.42 |

#### 2.2.2. Calculation of EEG signal features

Channels were grouped into frontal (F7, F3, Fz, F4, F8), central (Cz, C3, C4), and occipital (O1, O2) regions (Figure 1D). The power spectral density (PSD) of EEG signals was calculated using Welch’s method [62] (1.0s Hamming windows, 0.5s overlap, 0.1-50 Hz range). Spectral parameterization was limited to 0.5-13 Hz to minimize muscle artifacts [5], with parameters: peak width limits (1, 12.0), maximum peaks: 6, minimum peak amplitude: 0.0, peak threshold: 2.0, aperiodic mode: “fixed”. Including frequencies above 13 Hz systematically increased model error and biased aperiodic parameters by forcing the model to accommodate high frequency activity, which is particularly problematic in pediatric autism populations where movement artifacts (>13 Hz) are prevalent (Figure Supplement 2). Analysis yielded aperiodic exponent, offset, and aperiodic adjusted band powers (delta: 0.1-4 Hz, theta: 4-8 Hz, alpha: 8-13 Hz) (Figure 1E, Figure Supplement 1E). Channels with model fits R^2^ <0.80 were excluded. Aperiodic corrected oscillatory power was calculated by subtracting the aperiodic fit from the PSD and finding the maximum corrected power within each band, then averaging within channel group.

#### 2.2.3. Statistical analysis

Blood metabolite levels were z-scored to address non-normality from detection thresholds. Our dataset showed variability in sample sizes across assessments due to assessment administration difficulties, experimenter error, or low participant cooperation. To account for missing data, we used Linear Mixed Effects (LME) models which are advantageous for handling data where time points are nested within participants and can account for this type of data dependency, as used in other studies where children were able to complete certain assessments but not others [63]. Our statistical approach examined three distinct electrode groups (frontal, central, occipital) based on established functional neuroanatomy and connectivity patterns. For each electrode group, we analyzed relationships with three biochemically distinct CBD metabolites (CBD, 7-OH-CBD, 7-COOH-CBD), which have different pharmacological properties and half-lives. Each model included random subject intercepts and fixed effects for randomization, timepoint, age, ADOS-2 module used to assess participant as a proxy for participants’ language ability, and days since last blood extraction for metabolite measurements. Specifically, timepoint (Baseline, Post-placebo, Post-CBD, Post-wash) and Randomization order (CBD-first or Placebo-first) were included to account for potential practice effects from repeated measurements. Three metabolite models were applied to each electrode group (totaling 9 models) and cognitive-behavioral assessment (totaling 15 models). To further interpret our findings, we calculated the estimated marginal means (EMMs) for post-CBD timepoints derived from significant model coefficients and analyzed the relationship between EMMs and changes in post-CBD and baseline measures for participants that had data for both sessions included in the linear mixed model. The number of sessions included in each model is listed in Table 2. Post-hoc regression analyses using estimated marginal means were conducted to visualize the directionality and magnitude of significant mixed model relationships.

## 3. Results

### 3.1. Broadband and aperiodic adjusted task-free EEG features across the scalp had a positive association with CBD blood metabolite levels

We found a clear increase across all CBD blood metabolites levels following the 8-week CBD treatment period (Figure 1A, B). Prior EEG studies on intellectually disabled and minimally verbal neurodevelopmental disorder populations have shown heterogeneity in neural spectral power and aperiodic measures of scalp EEG activity grouped by frontal, central, and occipital electrodes [10,14,16]. We first confirmed that post-processed EEG durations were similar across study time points (Figure 1C) and used a similar approach by grouping channels into frontal, central, and occipital areas to investigate the relationship between periodic and aperiodic EEG measures and CBD treatment (Figure 1D, E).

Our linear mixed effect models confirmed that CBD treatment increased metabolite levels in blood (p’s < 0.001; Figure 2A, C, F). CBD metabolite levels had varying relationships with periodic and aperiodic measures of EEG across brain regions. In frontal electrodes, 7-COOH-CBD levels were associated with increased aperiodic offset (β = 0.090, 97.5% CI = [0.023, 0.158], p = 0.009; Figure 2A, bottom), as confirmed by post-hoc analysis of estimated marginal means from baseline to post-CBD (n = 20, R^2^ = 0.510, p < 0.001; Figure 2B). Central electrodes showed mixed effects, with higher CBD metabolite levels associated with decreased aperiodic adjusted alpha power (β = −0.135, 97.5% CI = [-0.267, −0.002], p = 0.046; Figure 2C, top; post-hoc analysis of n = 20 participants, R^2^ = 0.379, p = 0.004; Figure 2D), while 7-COOH-CBD metabolite levels were associated with an increased aperiodic offset (β = 0.094, 97.5% CI = [0.026, 0.161], p = 0.006; Figure 2C, bottom; post-hoc analysis of n = 20 participants, R^2^ = 0.495, p < 0.001; Figure 2E). In occipital electrodes, higher 7-COOH-CBD metabolite levels were associated with increased aperiodic offset (β = 0.099, 97.5% CI = [0.033, 0.165], p = 0.003; Figure 2F, bottom; post-hoc analysis of n = 20 participants, R^2^ = 0.476, p < 0.001; Figure 2G) and decreases in aperiodic exponent (β = −0.235, 97.5% CI = [-0.460, −0.010], p = 0.041; Figure 2F, bottom), though this latter relationship was relatively modest in post-hoc analysis (n = 20 participants, R^2^ = 0.058, p = 0.308; Figure 2H).

**Figure 2.**
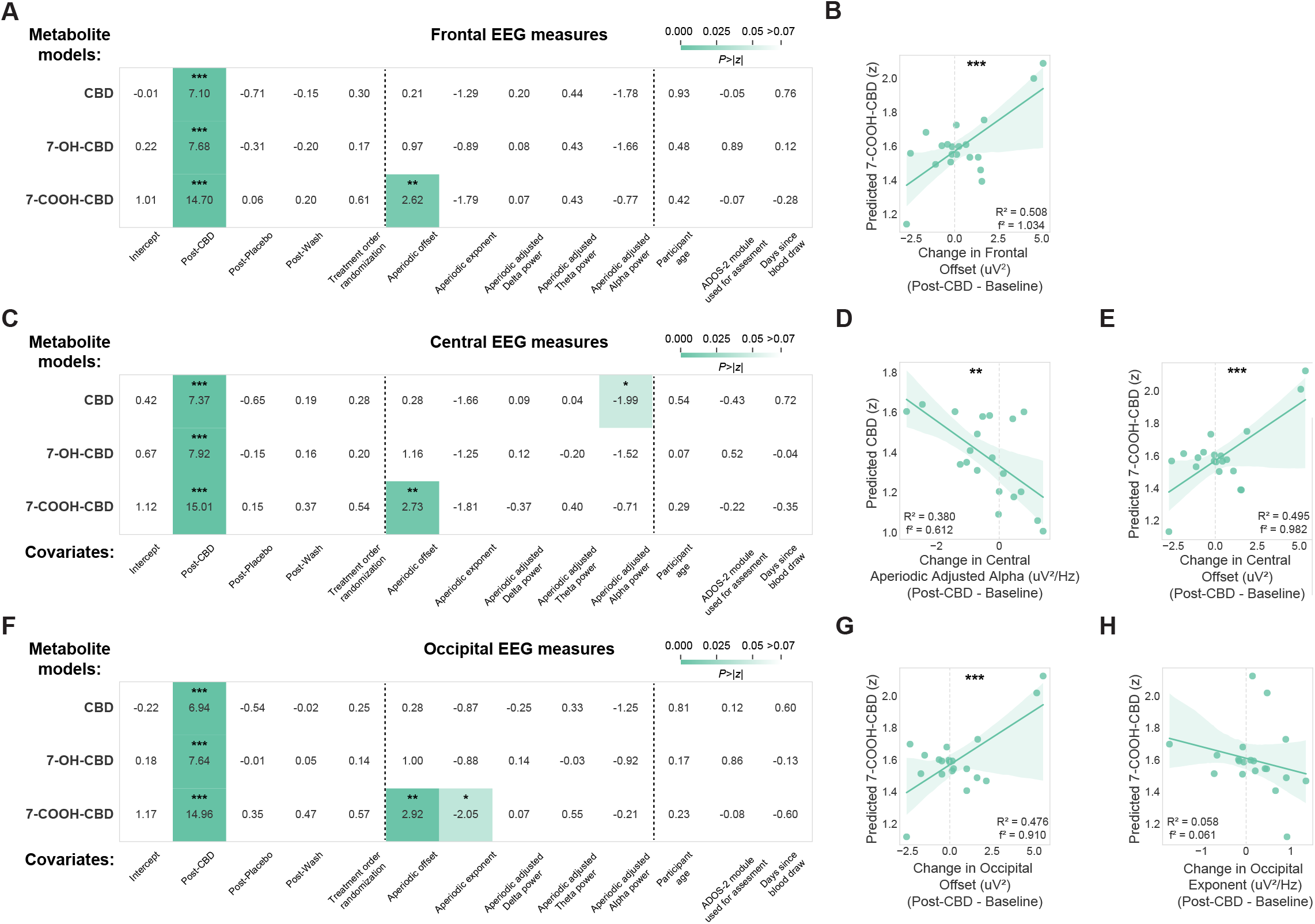
Relationships between task-free EEG power spectrum features and CBD blood metabolite levels. Results from linear mixed models and linear regressions relating periodic and aperiodic measures of EEG signals to levels of CBD, 7-OH-CBD, and 7-COOH-CBD metabolite in blood across groups of electrodes correspondingly located above **(A, B)** frontal, **(C–E)** central, and **(F–H)** occipital regions. Visualization of each linear mixed model shows the z-statistic for each model coefficient and a heatmap of the two-tailed p-value associated with the z-statistic. *p<0.05, **p<0.01, ***p<0.001.

### 3.2. Measures of receptive vocabulary, nonverbal intelligence, and visuomotor coordination had a positive association with CBD blood metabolite levels

Our linear mixed effect models showed a mixture of positive associations between CBD blood metabolite levels and cognitive-behavioral performance measures after controlling for age, ADOS-2 module, and treatment order. Higher CBD metabolite levels were associated with improved receptive vocabulary on the PPVT-4 (β = 0.004, 97.5% CI = [0, 0.007], p = 0.039; Figure 3A, top), though this relationship was modest in post-hoc analysis (n = 12 participants, R^2^ = 0.225, p = 0.119; Figure 3B). Similarly, increased 7-COOH-CBD levels were associated with higher nonverbal intelligence scores on the TONI-4 (β = 0.012, 97.5% CI = [0.005, 0.019], p < 0.001; Figure 3C, bottom), with post-hoc analysis showing a modest relationship (n = 10 participants, R^2^ = 0.010, p = 0.780; Figure 3D). The strongest CBD treatment improvements appeared in visuomotor integration abilities. Both 7-OH-CBD and 7-COOH-CBD blood metabolite levels were positively associated with Beery-Buktenica VMI test performance, with 7-OH-CBD showing a modest effect (β = 0.034, 97.5% CI = [0.002, 0.066], p = 0.038; Figure 3E, middle) with post-hoc analysis showing a modest relationship (n = 18 participants, R^2^ = 0.136, p = 0.131; Figure 3F). Beery-Buktenica VMI scores particularly improved with higher 7-COOH-CBD levels (β = 0.036, 97.5% CI = [0.018, 0.054], p < 0.001; Figure 3E, bottom), confirmed by post-hoc analysis (n = 18 participants, R^2^ = 0.257, p = 0.032; Figure 3G). ADOS-2 module used to assess participants showed a consistent negative relationship with 7-COOH-CBD blood metabolite levels across in the PPVT-4 (β = −0.194, 97.5% CI = [-0.384, −0.004], p = 0.046; Figure 3A, bottom) and TONI-4 (β = −0.192, 97.5% CI = [-0.372, −0.011], p = 0.037; Figure 3C, bottom) models. CBD treatment showed no significant effects on repetitive behaviors (RBS assessment) or expressive vocabulary (EOWPVT-4 assessment) (Figure Supplement 3).

**Figure 3.**
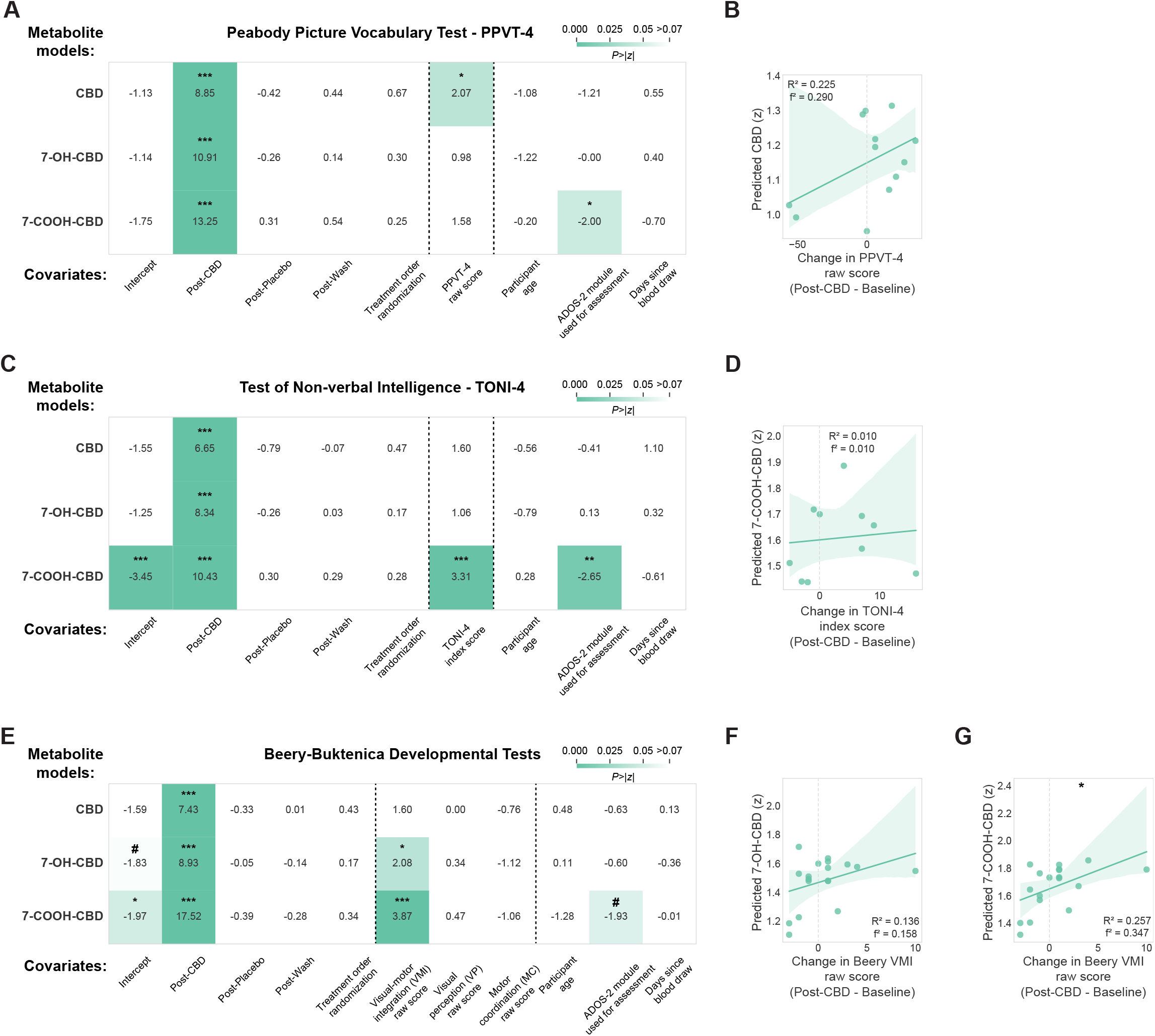
Relationships between cognitive-behavioral assessments and CBD blood metabolite levels. Results from linear mixed models and linear regressions relating **(A, B)** receptive vocabulary ability, **(C, D)** non-verbal intelligence, and **(E–G)** visuomotor coordination assessment scores to levels of CBD, 7-OH-CBD, and 7-COOH-CBD metabolite in blood. Visualization of each linear mixed model shows the z-statistic for each coefficient and a heatmap of the two-tailed p-value associated with the z-statistic. #p<0.06, *p<0.05, ***p<0.001.

## 4. Discussion

Our secondary analyses from a double-blind, placebo-controlled trial provided longitudinal measures of oral CBD effects on task-free scalp EEG signal features, CBD-related metabolism, and cognitive-behavioral assessments in children with ASD requiring higher levels of support. Improvements in visuomotor coordination, alongside more modest gains in nonverbal intelligence and receptive vocabulary abilities, but not in expressive language or repetitive behaviors, suggest CBD treatment and related metabolites may differentially affect distinct aspects of brain function. The strongest associations were observed in visuomotor tasks, potentially stemming from CBD’s modulation of GABAergic signaling in brain regions critical for motor control and visual processing [40,64,65]. The concurrent improvements in receptive vocabulary and nonverbal intelligence, although more modest, align with previous CBD:THC preparation studies that reported improvements in social and cognitive deficits in ASD patients [35,36]. While group-level analyses showed positive trends for these measures, individual participants showed varying magnitudes of change, with some demonstrating robust improvements while others showed minimal or negative changes in relation to CBD metabolite levels, highlighting the importance of considering individual variability in treatment responses.

Our EEG findings provide insight into CBD treatment’s potential impact on neural function. Specifically, the predominance of aperiodic over periodic effects suggests CBD treatment influences brain activity through changes in basic cellular and network properties rather than through specific excitatory or inhibitory circuits that mediate oscillatory activity. Aperiodic measures of brain activity are thought to reflect changes in broadband power as a function of neuronal population spiking [66,67]. This broadband change in neural activity aligns with magnetic resonance spectroscopy evidence showing CBD’s broad modulation of glutamate-GABA systems in ASD patient cortex [40]. The negative relationship between CBD metabolite levels and the aperiodic exponent in occipital regions may reflect CBD’s reported decrease of GABA+ macromolecules in ASD, as reduced inhibitory signaling typically decreases the aperiodic exponent. This relationship could potentially explain the differential effects on behavioral measures, with tasks requiring extensive visuomotor integration showing the strongest positive association with CBD blood metabolite levels, possibly due to their reliance on GABAergic signaling in visual and motor circuits. The inverse relationship between CBD metabolite levels and aperiodic-adjusted alpha power in central regions might similarly indicate a broad shift toward increased cortical excitability rather than inhibition-mediated oscillatory signaling [40]. The robust spatial consistency of changes in aperiodic features across the scalp, combined with their established test-retest reliability in ASD populations [8], suggests these measures may provide complementary EEG biomarkers for tracking treatment outcomes alongside traditional oscillatory measures, whose associations with CBD treatment were observed to be more spatially restricted.

Given that aperiodic measures are increasingly recognized as indicators of neurotypical brain development, their alterations in relation to CBD blood metabolite levels here suggest their potential utility as biomarkers for treatment responses in ASD, further extending previous work showing EEG features can serve as reliable biomarkers in human neurodevelopmental disorders [10,14–16]. Specifically, we observed a negative relationship between levels of 7-COOH-CBD metabolite in blood and the aperiodic exponent in occipital regions, indicating a decrease of the aperiodic exponent, as reflected by a “flatter” slope of the power spectrum model fit after CBD treatment, which is observed in typically developing children [59,68,69]. In contrast, we saw a positive relationship between levels of 7-COOH-CBD metabolite in blood and the aperiodic offset, indicating an increase in broadband power that contrasts with the decreasing patterns seen in typical neurodevelopment [68,69]. Our findings suggest that this CBD regimen may not induce all the expected trajectories of aperiodic activity throughout neurodevelopment, mirroring how the treatment regimen did not improve all cognitive-behavioral measures and had individual participant variability in responses.

Our restriction of spectral parameterization to 0.5-13 Hz aligns with established autism EEG biomarker research, where peak alpha frequency (6-12 Hz) correlates with non-verbal cognition and delta rhythms (1-4 Hz) show characteristic disruptions [16,24,25]. While this range excluded potentially relevant 25-35 Hz beta oscillations (also termed “iota”) that have been reported in healthy children and those with Fragile X syndrome [7,70], our focus on lower frequencies aimed to capture the established biomarkers while maintaining interpretability of aperiodic parameters as indices of E:I balance.

This restriction also avoided distinguishing genuine high frequency oscillations from movement artifacts in our behaviorally challenging population. Future studies could investigate CBD’s effects on higher frequencies, but for our research question about established lower-frequency markers of autism, the 0.5-13 Hz range provided robust model fits (mean R^2^ = 0.987) while avoiding interpretational ambiguities.

Our treatment regimen was derived from prior CBD trials in epilepsy, but the optimal duration for treatment effects likely differs [29,30]. The relationship between CBD metabolite levels in blood and cognitive-behavioral outcomes is notable, particularly given that the most consistent effects were observed with 7-COOH-CBD, which doesn’t cross the blood-brain barrier. This suggests that the effects we observed may involve indirect peripheral mechanisms in addition to direct neural modulation by CBD and 7-OH-CBD metabolites. Whether similar cognitive benefits would emerge with different treatment durations (either shorter or longer than 8 weeks) remains an open question warranting future studies, as we are not aware of any CBD-related clinical trials that have specifically examined these cognitive-behavioral measures. Longer-term follow-up is necessary to determine if the observed cognitive improvements persist after treatment discontinuation. The magnitude of effects we observed on visuomotor function compared to other domains may reflect neural circuit-specific responses to CBD, but it could also reflect differential test sensitivity to change, or general attentional improvements. The Beery VMI requires only drawing one additional line correctly for improvement, whereas language measures require learning new vocabulary over 8 weeks, which is a much more demanding change for children with neurodevelopmental conditions. Additionally, if CBD enhances attention and focus, this could more readily improve visuomotor tasks (where attention to detail directly improves performance) than vocabulary tasks (where attention cannot compensate for unknown words). Findings from the primary clinical trial support this interpretation, such that blinded clinicians observed reduced aggression, hyperactivity, and anxiety in 68% of children during CBD treatment, with many showing calmer behavior and improved communication [39]. These behavioral improvements suggest that CBD’s cognitive effects may be mediated through multiple mechanisms, including both enhanced attention and reduced disruptive behaviors during testing, as well as potential direct domain-specific neural changes, which could all contribute to the observed improvements.

Our results revealed substantial individual variability in treatment responses in relation to CBD blood metabolite levels. While group-level analyses showed positive associations, individual participants showed varying magnitudes of change, with some reflecting no change or even decreases in outcome measures. This heterogeneity likely reflects the complex pharmacology of CBD metabolism. The inactive 7-COOH-CBD metabolite has a longer half-life (>48 hours) compared to CBD and 7-OH-CBD, which decrease rapidly after 2-8 hours post-administration [48]. Since assessments weren’t consistently conducted within 8-12 hours of blood draws, measuring 7-COOH-CBD provided more reliable treatment indicators than CBD or 7-OH-CBD levels alone. We observed robust relationships between 7-COOH-CBD levels and both EEG features and cognitive-behavioral measures, despite this metabolite’s inability to cross the blood-brain barrier, suggesting CBD’s therapeutic effects involve both direct neural modulation via CBD or 7-OH-CBD and indirect peripheral mechanisms. For future studies, we recommend conducting blood draws shortly before assessments to better capture CBD and 7-OH-CBD effects. The absence of preserved weight data limited our reporting to weight-based dosing protocols, so future work should document both weight-based and absolute dosing metrics to provide a more precise measure of the actual amount of medication administered while incorporating longer intervention periods with validated measures for detecting short-term changes. Additionally, recent evidence of sex-based differences in 7-COOH-CBD metabolism [46] suggests that understanding metabolite relationships could help explain outcome variability and inform personalized dosing strategies, particularly in clinical settings where precise timing is challenging with pediatric ASD populations.

Conducting longitudinal studies in ASD children with higher support needs presented challenges that required flexibility in EEG data collection approaches. In our study, children viewed self-selected videos during EEG recordings due to the challenge of keeping them engaged, and differences in video content were not documented, leaving potential video stimulus-related effects on brain activity unknown. However, prior work with similar populations suggests this passive viewing approach does not confound interpretations of holistic neural activity measures [10]. This approach represented a necessary compromise that enabled the study of neural activity in this historically understudied population. Furthermore, the detection of scalp-wide aperiodic measure effects, which tend to be stable across behavioral conditions in pediatric and ASD populations [5,8], suggests robust, global changes in neural activity rather than behavioral state artifacts that might be induced by specific video content. In addition, our findings are limited to male children, which limits generalizability to females with ASD. This was an intentional design choice in the original study, given the small sample size recruited and known differences in autism symptomatology between males and females that may have confounded findings [39].

A key limitation in interpreting our results stems from the scarcity of research validating cognitive and behavioral test sensitivity within brief intervention periods, particularly for children with autism who have higher support needs. To our knowledge, only one study has examined short-term changes in TONI-4 performance, finding small but detectable changes in adult schizophrenia patients over just a two-week period [71]. While we observed improvements in visuomotor coordination, receptive vocabulary and nonverbal intelligence measures in relation to CBD blood metabolite levels, the clinical meaningfulness of these changes requires careful consideration. An additional limitation is that while our linear mixed effects models detected significant associations when controlling for multiple covariates, several relationships showed modest effect sizes in post-hoc linear regressions, including receptive vocabulary (PPVT-4), nonverbal intelligence (TONI-4), and some EEG measures (aperiodic exponent in occipital electrodes). It is possible that these represent subtle effects that emerge primarily through rigorous statistical control rather than strong standalone associations, requiring cautious interpretation and replication in larger samples. The use of raw scores for EOWPVT-4, PPVT-4, and Beery VMI-6 assessments rather than standardized scores helped avoid floor effects and maintain sensitivity to detect changes, but this approach also makes it more challenging to contextualize the magnitude of improvements against established clinical thresholds. Furthermore, individual variability in our results likely reflects both genuine treatment response differences and measurement unreliability from variable attention and cooperation during testing, with lower scores being more susceptible to such confounds than higher scores. Additionally, the 8-week intervention period, while consistent with other pharmacological trials, may not have been optimal for capturing the full extent of potential cognitive changes, as developmental improvements often unfold over longer timeframes. Our CBD metabolite levels aligned with previous oral administration studies in healthy adults [41] and a child with Dravet Syndrome [72], though age-related metabolic differences should be considered when interpreting these comparisons Developing novel ASD treatments requires reliable methods to characterize associated brain activity changes. While task-free EEG measurements have shown promise as biomarkers of ASD severity, even from 3 months of age [24,73–75], these studies did not separate periodic and aperiodic components of neural power spectra, obscuring the specific neural processes affected. More recent work shows age-dependent variations: adult ASD populations show mixed differences in task-free EEG features [24,76], while in preterm children, aperiodic features are associated with increased ASD risk [77]. Notably, aperiodic measures of EEG activity in 6-8-year-old children with ASD demonstrates high test-retest reliability compared to healthy controls [8], supporting its potential as a developmental biomarker. Our study addresses these gaps by applying spectral parameterization to EEG recordings from near-adolescent ASD children with well-defined higher support needs.

In conclusion, we report that metabolic responses to an 8-week daily CBD regimen using a purified formulation with regulatory precedent were associated with improvements to select domains of cognitive and behavioral ability, such as visuomotor coordination, receptive vocabulary ability and nonverbal intelligence, and that these improvements coincided with neural activity changes primarily in the aperiodic component of task-free scalp EEG signals.

## Data Availability Statement

The data presented in this study will be available on request once data analyses are completed from UC San Diego Center for Medicinal Cannabis Research (CMCR) through contacting Doris Trauner, M.D. The EEG and cognitive-behavioral assessment data are not publicly available to protect the children’s and their families’ privacy. The analysis code needed to reproduce the analysis and figures is provided here: https://github.com/voytekresearch/cbd-asd-eeg

## Acknowledgements

We’d like to thank Dillan Cellier, Dr. Sydney Smith, and Dr. Maribel Patiño for their thoughtful discussion.

## Author Contributions

C.C.: Formal analysis, Investigation, Methodology, Visualization, Writing - original draft, Writing - review and editing.

G.P.: Methodology, Data acquisition, Writing - review and editing.

A.H.: Writing - review and editing.

D.T.: Conceptualization, Project administration, Funding acquisition, Investigation and Writing - review and editing.

B.V.: Supervision, Project administration, Funding acquisition, and Writing - review and editing.

---

**Figure Supplement 1.**
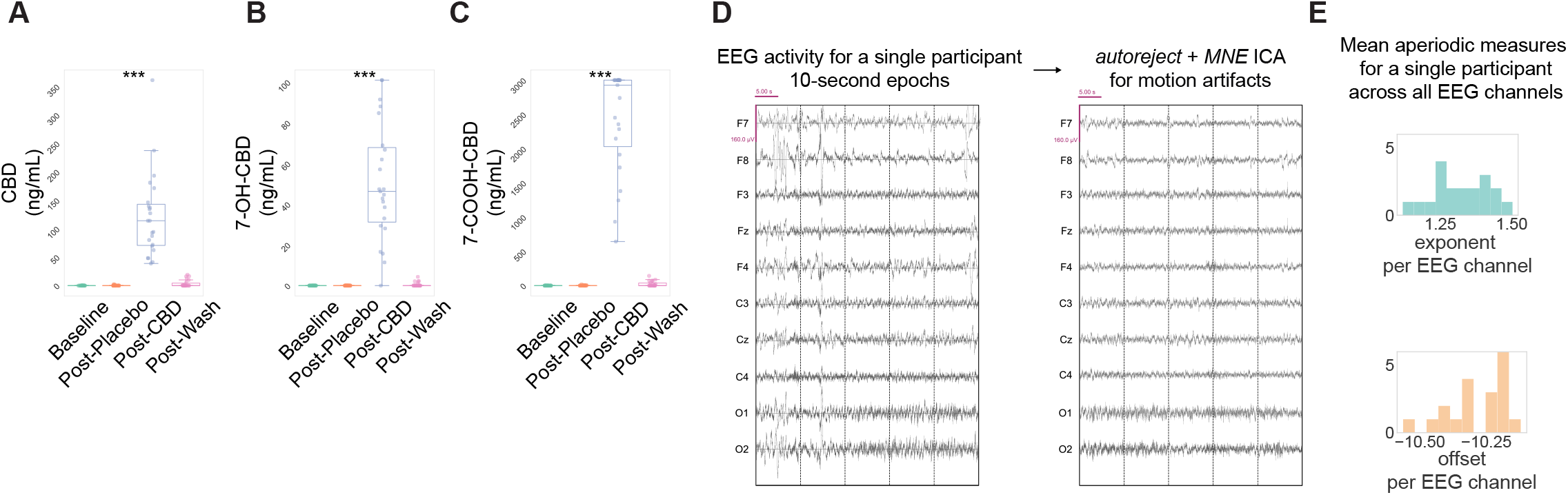
CBD blood metabolite levels and preprocessing of task-free EEG signals. **(A)** Concentration of metabolite levels in blood, including CBD, **(B)** 7-OH-CBD and **(C)** 7-COOH-CBD, across all study timepoints. **(D)** Representative traces of segmented EEG before and after artifact rejection. **(E)** Representative aperiodic exponent and offset values for all EEG electrode channels in a single session for a single participant.

**Figure Supplement 2.**
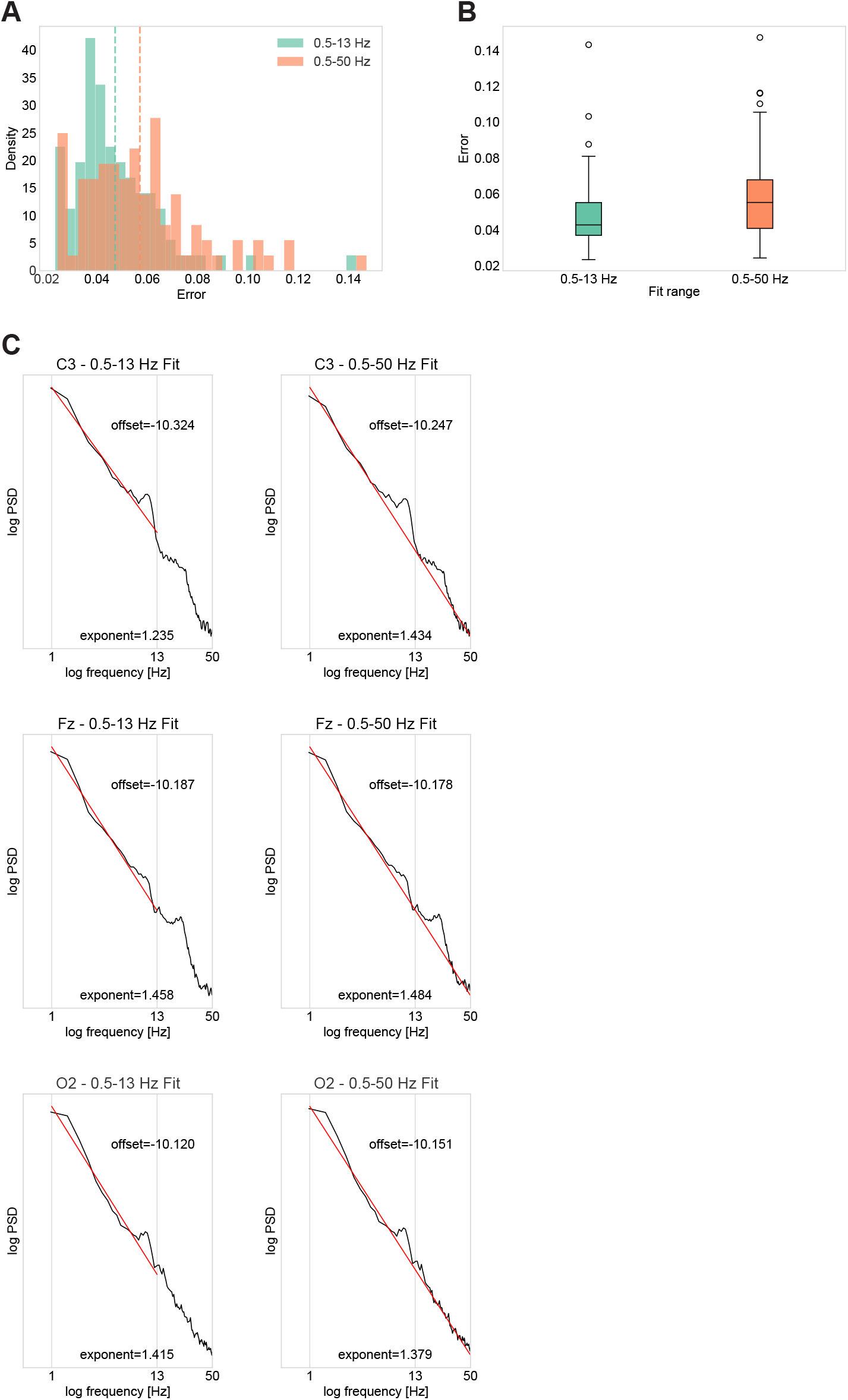
Spectral parameterization frequency range comparison. **(A)** Error distribution comparison across all channels shows that 0.5-13 Hz fits (teal) produce systematically lower model errors compared to 0.5-50 Hz fits (orange), with means of ∼0.04 and ∼0.06 respectively. (B) Box plot comparison showing the consistent reduction in model error with 0.5-13 Hz frequency restriction. (C) Representative examples demonstrate how 0.5-50 Hz fits (right column) attempt to model high-frequency artifacts, resulting in high frequency noise-biased aperiodic parameters compared to the 0.5-13 Hz fits (left column).

**Figure Supplement 3.**
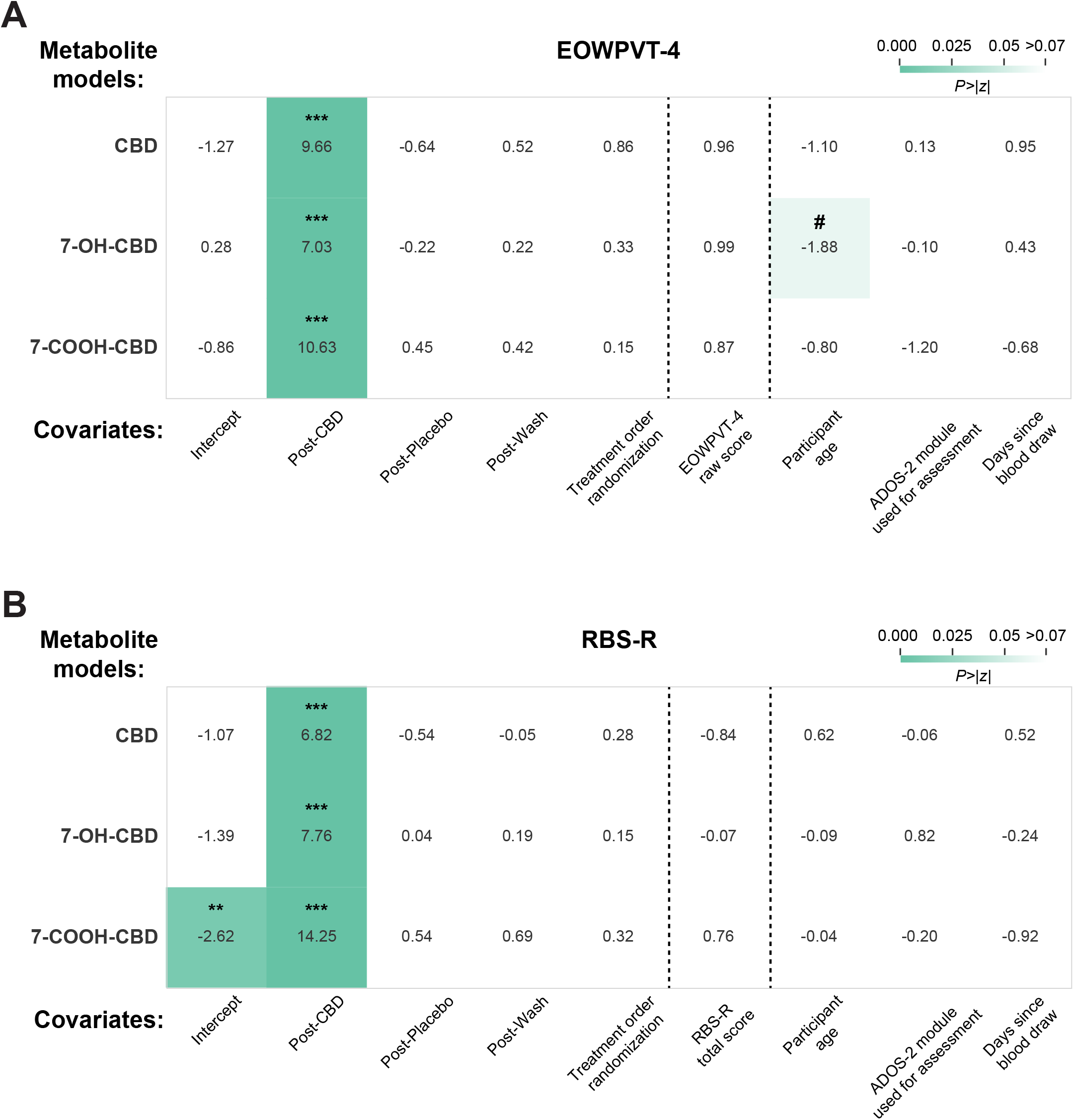
Absent relationships between cognitive-behavioral assessments and CBD blood metabolite levels. Results from linear mixed models and linear regressions relating (**A)** repetitive behavior, **(B)** expressive vocabulary assessment scores to levels of CBD, 7-OH-CBD, and 7-COOH-CBD metabolite in blood. Visualization of each linear mixed model shows the z-statistic for each coefficient and a heatmap of the two-tailed p-value associated with the z-statistic. #p<0.06.

